# Disentangling Perinatal Depression from General Major Depression: A GWAS Meta-Analysis and Genetic Correlation Approach in European Datasets

**DOI:** 10.1101/2025.11.23.25340751

**Authors:** Jared G. Maina, Jerry Guintivano, Rujia Wang, Laura Meldrum, Saakshi Kakar, Danyang Li, Sang-Hyuck Lee, Helena L. Davies, Gursharan Kalsi, Anthony J. Cleare, Katrina A.S. Davis, James T.R. Walters, Daniel J. Smith, Chérie Armour, Andrew McIntosh, Nathalie Kingston, John R. Bradley, The GLAD Study, NIHR BioResource Consortium, Matthew Hotopf, Evangelos Vassos, Jonathan R.I Coleman, Thalia Eley, Gerome Breen

## Abstract

Perinatal depression (PND), defined as depression occurring during the perinatal period, has a higher heritability than general major depressive disorder (MDD) yet remains understudied. This study aimed to increase power in PND GWAS and explore its genetic architecture. We meta-analysed an existing study by the Psychiatric Genomics Consortium (PGC) with data from the GLAD+ study and UK Biobank (UKB), yielding 25,452 PND cases and 72,131 controls. Linkage disequilibrium score (LDSC) regression was used to compare genetic correlation profiles of PND and general MDD across traits. Gene, gene-set, and tissue expression analyses were also performed. The genetic correlation between PGC PND and UKB-GLAD+ PND was high (0.93). No genome-wide significant loci emerged from the meta-analysis. The SNP-based heritability of PND on the liability scale was 0.089. PND and MDD displayed broadly similar genetic correlation patterns. Still, PND had a significantly higher genetic correlation with anxiety disorders, while MDD showed higher correlations with cannabis use disorder, bipolar disorder, and ADHD. Tissue expression analysis showed the highest enrichment in brain regions like the cortex and nucleus accumbens. The sample size was not large enough to detect genome-wide significant loci, and further analyses in ancestrally diverse datasets are needed. PND definitions were heterogeneous, and precise information on onset timing across studies was lacking. This study, the largest European ancestry meta-analysis of PND to date, suggests distinct genetic factors may differentiate PND from MDD in general. The findings highlight the need for larger and more ancestrally diverse studies to identify genome-wide significant loci.

## INTRODUCTION

The perinatal period is associated with increased risk of psychiatric conditions, the most common of which is perinatal depression (PND), with a lifetime prevalence estimate of 10%(1). PND encompasses depression arising during pregnancy (antenatal depression) or shortly after childbirth (postnatal depression). The prevalence ranges from 10-25%(1), with a higher prevalence in low- and middle-income countries than in high-income countries(1,2). Twin studies suggest PND is a more heritable subtype of major depressive disorder (MDD) (54% for PND versus 37% for general MDD)(3). Despite its higher heritability, PND is understudied. The latest PND GWAS led by the Psychiatric Genomics Consortium (PGC) was underpowered to detect genome-wide signals, suggesting larger sample sizes are required to increase power.

In this present study, we aimed to increase power in PND GWAS by combining existing published studies with data from the Genetic Links to Anxiety and Depression Study and other UK National Institute of Health Research (NIHR) BioResource cohorts (GLAD+)(4) and the UK Biobank (UKB)(5). We then explored the genetic architecture of PND by applying linkage disequilibrium score (LDSC) regression(6) to compute genetic correlations between PND and MDD in general, as well as other phenotypes.

## MATERIALS AND METHODS

### Study datasets

In total, this study included 25,452 cases and 72,131 controls (total N = 97,583) by combining data from the UK Biobank and the GLAD+ with the recent GWAS of PND led by the PGC (**Table 1**).

**Table 1.**
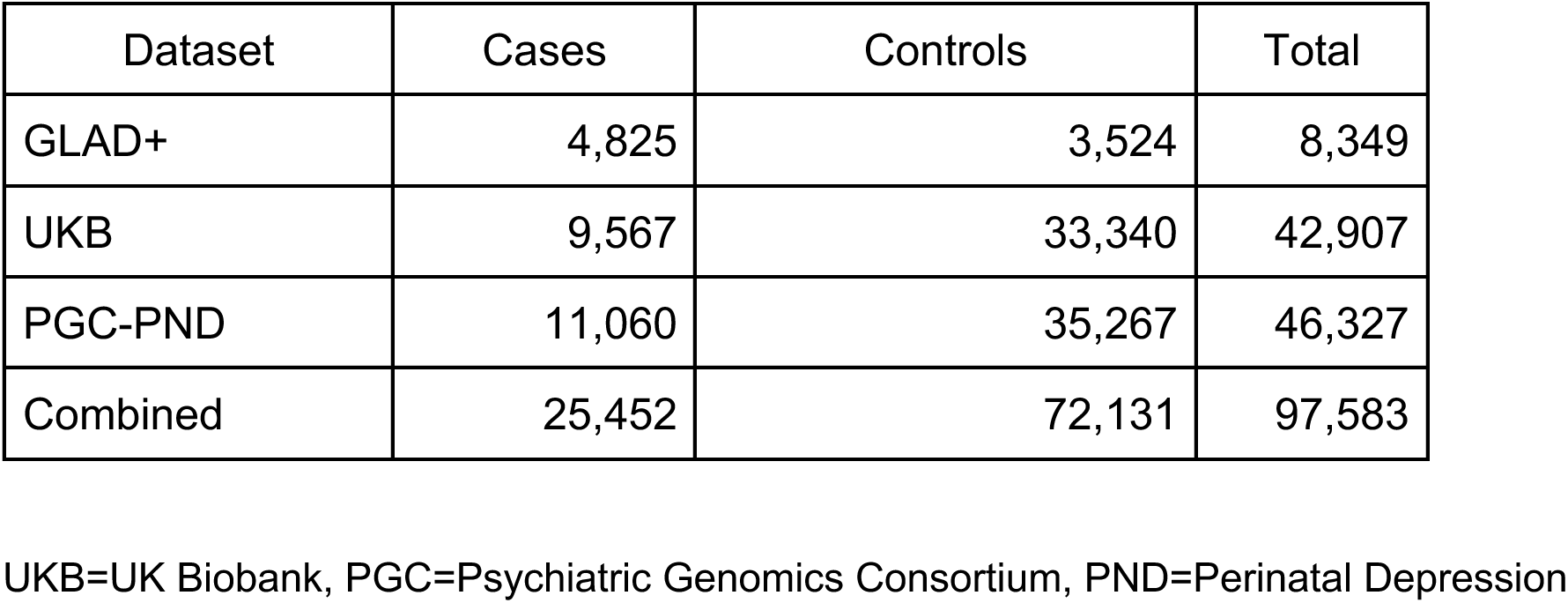
Study numbers of different study datasets included in the meta-analyses.

#### GLAD+

The GLAD+ consisted of participants recruited from two UK studies: the GLAD Study (www.gladstudy.org.uk), and the NIHR BioResource (NBR) COVID-19 Psychiatry and Neurological Genetics (COPING) study(7). The GLAD Study is an ongoing study of anxiety and depression in the UK with more than 70,000 consented participants recruited through social media campaigns and referral from clinical sites. Participants provide demographic and environmental information, and consent to medical record linkage and recontact for future research. Over 37,000 of these participants have completed the online recruitment survey and returned a saliva sample. Recruitment for the COPING study during the COVID-19 pandemic was carried out through email invitations sent to participants in the GLAD Study and to individuals in the broader NBR resource, which includes members of an Inflammatory Bowel Disease cohort as well as general population cohorts. Interested individuals enrolled by following a study link, providing informed consent online, and completing a digital questionnaire. All GLAD and NBR study participants were eligible to take part. GLAD+ consisted of cases from the GLAD study, while controls were healthy volunteers from the COPING study.

#### UKB

We used data from the UKB (https://www.ukbiobank.ac.uk), which consists of approximately 500,000 participants recruited across 22 centres in the UK between 2006-2010. Participants in the UKB provided information through baseline questionnaire assessment, linkage with hospital inpatient records and targeted follow-up questionnaires such as the mental health questionnaires. The first mental health questionnaire (MHQ1)(8) was rolled out in 2016-2017 and received responses from over 157,000 and the second mental health questionnaire (MHQ2) was sent to participants in 2022, leading to some new respondents, and 215,250 participants with questionnaire data in total(9).

#### PGC PND summary statistics

This analysis incorporated the most recent GWAS summary statistics for PND from a large-scale study of European samples led by the PGC. The details of the individual studies included in the PGC meta-analyses have been elaborated elsewhere(10). In their analyses, the PGC only included data from the MHQ1 in UKB. To avoid sample overlap in our UKB analyses, we used leave-one-out summary statistics from the PGC dataset that excluded UKB participants.

### PND Definition

In GLAD+, PND cases were defined using two approaches:

1. Self-reported perinatal depression definition. Participants were asked: “*Have you ever been diagnosed with one or more of the following mental health problems by a professional, even if you don’t have it currently?*” Female participants who selected “*Depression during or after pregnancy*” were classified as PND cases.
2. Composite International Diagnostic Interview (CIDI) for Depression (CIDID) algorithm-based definition. Female participants were classified as PND cases if they responded “*Yes*” to the question: “*Did any of these symptoms (Low mood for 2 weeks or anhedonia for 2 weeks) occur within the first year of giving birth? Or has it been suggested you had post-natal depression*”.

After deriving PND cases using both approaches, we combined all female participants classified as cases by the CIDID algorithm with those not classified as CIDID cases, but who self-reported a PND diagnosis. Thus, a “Yes” response for either definition was sufficient for case inclusion. PND controls were women with no history of MDD, as defined by the COPING study criteria. In COPING, MDD was assessed with the adapted CIDI–Short Form (CIDI-SF). Participants were classified as controls if they did not meet Diagnostic and Statistical Manual of Mental Disorders Fifth Edition (DSM-5) criteria for MDD. These criteria required at least one core symptom (depressed mood or loss of interest) occurring daily or nearly daily for two weeks or more, together with five or more symptoms in total, including sleep disturbance, changes in weight or appetite, fatigue, impaired concentration, feelings of worthlessness, or suicidal ideation. In total, there were 8,349 individuals available for analyses in GLAD+ (4,825 cases and 3,524 controls) (**Table 1**). The median age of the GLAD+ participants included in this study was 51 years (interquartile range, 40 to 61).

In the UKB, the PND case control definition criteria applied by Guintivano et al. was employed, as described in detail elsewhere(10). In brief, PND cases were defined as women with at least one live birth who self-reported perinatal depression in either the Mental Health Questionnaire (MHQ) or the nurse-led interview. Controls were women with at least one live birth and no history of major depressive disorder (MDD). Further details on case and control definitions in UKB are provided in the **Supplementary Methods**. In total, we included 42,907 participants in UKB (9,567 PND cases, 33,340 as controls) (**Table 1**). The median age among UKB participants included in this study was 58 (interquartile range, 51-63).

The PGC PND summary statistics incorporated 18 European cohorts that used different phenotype definitions to ascertain PND cases, as described in their manuscript(10). Overall, the Edinburgh Postnatal Depression Scale (EPDS)(11), structured clinical interviews, and self-report assessments were used to define PND cases across these cohorts. The EPDS was the most frequently used tool among the studies included in the meta-analysis. The EPDS assesses current depressive symptoms using ten Likert-scale items, each rated from zero to three. In addition, the lifetime version of the EPDS was used to measure the lifetime occurrence of PND. PND symptoms are then rated on a scale of 0–30, with higher scores indicating greater symptom severity. Cases were defined if individuals had scores ≥13. Further details on the additional assessment instruments used across cohorts are available in the referenced manuscript(10). The summary statistics with UKB excluded (leave-one-out) included 46,327 individuals (11,060 cases and 35,267 controls) (**Table 1**) representing the combined sample from all other contributing cohorts.

### Genotyping, quality control and imputation

GLAD+ participants were genotyped using the UK Biobank Axiom Array v1 and v2 across multiple batches. Individuals of European ancestry were identified using GenoPred pipeline(12) by projecting onto principal components of genome-wide genotypes from 1000 Genomes(13). Quality control excluded variants with MAF < 0.01, call rate < 0.95, or HWE *P* < 1×10⁻⁸, and individuals with sex mismatches, heterozygosity outliers, sample duplications, or excess relatedness. After quality control, 33,635 individuals and 484,182 variants remained for imputation. Data were imputed to the TOPMed Freeze 8 reference panel(14) and filtered to retain variants with MAF ≥ 0.01 and imputation R² ≥ 0.3, yielding 15,009,228 variants for analysis. UKB genotyping and quality control have been described in detail elsewhere(5). In brief, participants were genotyped using the UK Biobank Axiom Array (90% of participants) or the UK BiLEVE array. Quality control excluded individuals with high missingness, heterozygosity outliers, or sex mismatches, and one from each related pair (KING > 0.04). Imputed genotypes for 487,422 individuals were based on 670,739 markers using IMPUTE4 and the Haplotype Reference Consortium (HRC)(15) and UK10K reference panels(16). SNPs with MAF < 0.01 or INFO < 0.4 were excluded. Post-QC, 413,259 participants remained, including 397,319 of European ancestry, identified via k-means clustering on principal components.

### UKB GLAD+ merged data

To maximise power in the UKB and GLAD+ case-control analyses, and with permission from UK Biobank and the NIHR, we merged UKB and GLAD+ participants into a single sample, combining individual-level hard-called imputed genetic data with the genotype hard-calling likelihood higher than 0.9 using Plink 1.9(17). We then performed a quality control on the combined data, restricting to variants with MAF > 0.01, missing rate < 0.02, Hardy-Weinberg equilibrium *P* > 1×10^−8^, and individuals’ missingness < 0.02. We assessed individual relatedness between UKB and GLAD+ participants using KING software(18). A total of 1,815 participant pairs shared identical genetic data; we assumed that pairs within the same dataset represent twins, while pairs across the two datasets represent duplicates (n = 1,633). Where a participant had provided genetic data for both studies (‘duplicates’), we preferentially retained the higher-quality UKB blood-based genotype data. Principal components (PCs) were projected onto the whole pooled sample from unrelated individuals using flashpca2(19). The resulting sample was 488,349 individuals and 8,274,311 imputed SNPs.

### GWAS and GWAS meta-analyses

We performed GWAS in the merged UKB and GLAD+ using the REGENIE software(20) (See **Supplementary Materials** for parameters used). We included the genotyping batch and the first 10 PCs as covariates. The statistical threshold for genome-wide significant SNPs used was *P* < 5×10^−8^.

METAL(21) was used to conduct a fixed-effect meta-analysis between PGC PND summary statistics and the merged UKB-GLAD+ summary statistics in 8,937,862 SNPs (See **Supplementary Material** for parameters used). Manhattan and QQ plots were generated using the *qqman* R package(22), while regional association plots were generated using FUMA version 1.7(23).

### SNP heritability and genetic correlation estimation

SNP-based heritability (h^2^_SNP_) was estimated on the liability scale in our analyses using precomputed LD score references provided by the LDSC software(6). We used a conservative 10% lifetime prevalence of PND when converting h^2^_SNP_ to the liability scale, consistent with the previous GWAS by Guintivano et al.(10). Genetic correlations between the PND results and a broad set of psychiatric disorders, sleep traits, reproductive traits, and behavioural phenotypes were estimated using LDSC. To assess how these correlations compared to those for general MDD, we used summary statistics from the latest PGC MDD study, excluding the 23andMe dataset(24). By comparing the genetic correlation profiles of PND and MDD across traits, we aimed to evaluate the degree of overlap and potential differences in their underlying genetic architectures. Statistical significance for the difference between the PND and MDD correlations was determined using the formula *P* = 2 × pnorm(–|Z|), where pnorm represents the cumulative distribution function of the standard normal distribution. The Z statistic was computed as 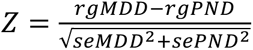, assuming a mean of 0 and a standard deviation of 1, as described previously(25). After Bonferroni correction for 22 genetic correlation tests, the adjusted significance threshold was set at *P* = 2.27 × 10^−3^.

### Gene-based, gene set and tissue expression analysis

To provide biological insight into the underlying biology of the meta-analyses results, we functionally mapped and annotated genetic associations from the meta-analysis using FUMA version 1.5.2((23)) (See **Supplementary Material** for parameters used). Gene and gene-set analyses were conducted using MAGMA version 1.08(26), as implemented in the FUMA platform. In the gene-based analysis, genome-wide SNPs were mapped to protein-coding genes in MAGMA. A competitive test was conducted for gene set analyses using default gene sets in FUMA from MsigDB version 7.0, totalling 15,496 gene sets (5,500 curated gene sets and 9,996 Gene Ontology terms). Curated gene sets were derived from nine data resources, including KEGG, Reactome, and BioCarta. Gene Ontology (GO) terms were grouped into three categories: biological processes, cellular components, and molecular functions. Gene expression datasets were obtained from GTEx version 8. MAGMA gene-property analyses were conducted using average gene expression per category (e.g., tissue type or developmental stage), conditioning on average expression across all categories in a one-sided test. This approach evaluates whether higher expression in a specific category is positively associated with genetic association signals.

## RESULTS

### GWAS and GWAS meta-analysis

The genetic correlation between PGC PND and UKB-GLAD+ PND was 0.93 (SE=0.12, Z=7.8, *P* = 4.5×10^−15^), indicating a high degree of concordance between the two studies for meta-analysis. After meta-analysis, no SNPs reached genome-wide significance (*P* < 5×10^−8^) for PND. However, 42 variants showed *P* < 1×10⁻⁵, a threshold often used to highlight potential exploratory signals. (**Figure 1, Supplementary Table 2**).

**Figure 1.**
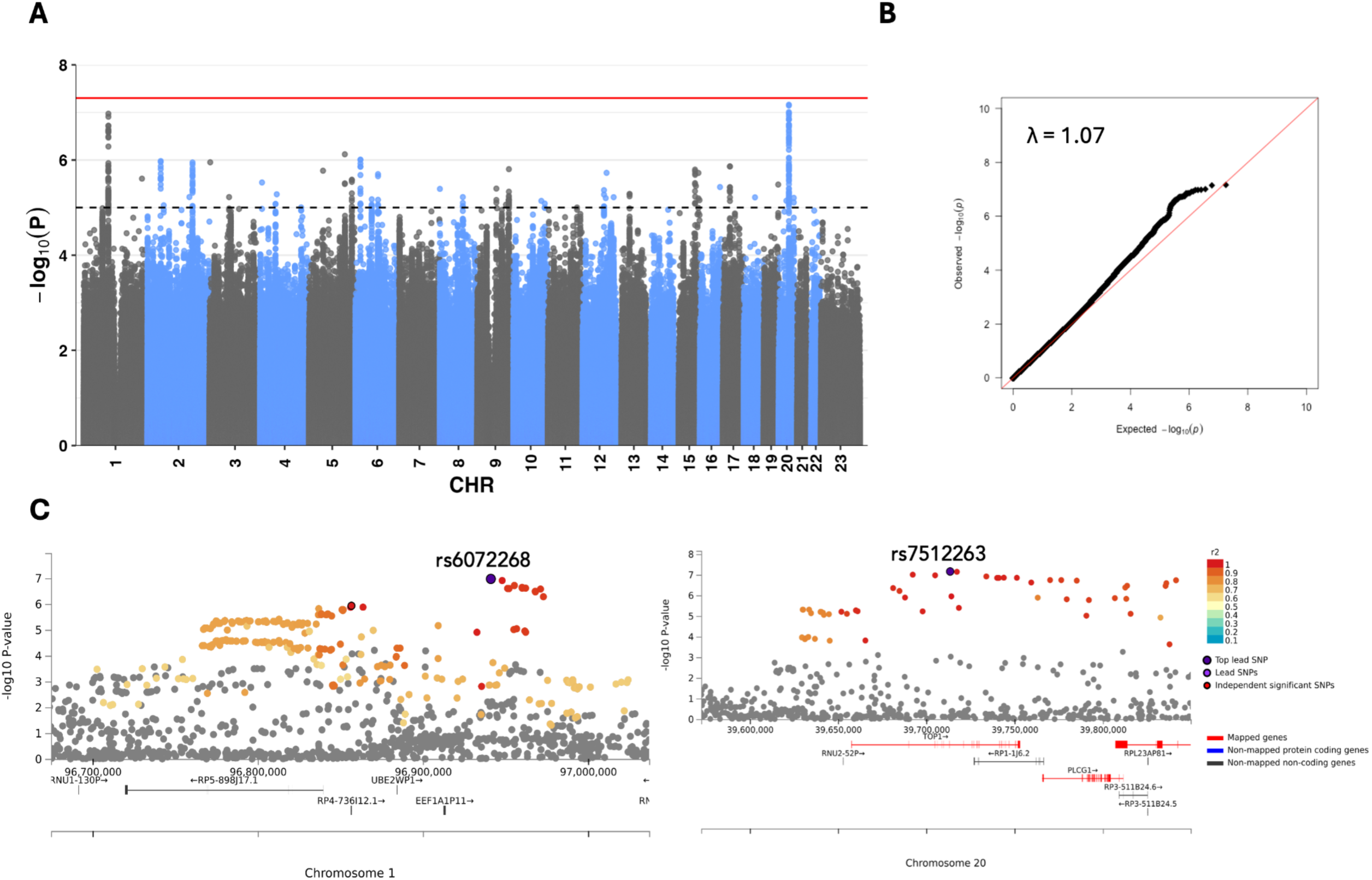
Manhattan, Quantile-Quantile (QQ) and regional plots of the PND meta-analysis results. **A**. Manhattan plot of the PND associations. The red solid line indicates the genome-wide significance threshold of *P* = 5×10^−8^, and the black dashed line represents a threshold of *P* = 1×10^−5^. **B.** QQ plot of association, λ=1.07. **C.** Regional plots of the top two loci on chromosomes 1 (left plot) and 20 (right plot)

The most significant SNP, rs6072268, is located on chromosome 20 (*P* = 6.89×10^−8^). This intronic SNP lies within the *TOP1* gene and close to the *RP1-1J6.2* gene (**Figure 1B**, **Table 2**). The same top SNPs were identified in the previous GWAS of PND (10), though our study showed marginally improved power. These top PND SNPs were also present in the recent MDD GWAS(24), but none reached significance in MDD; only one variant showed a marginal signal (*P* = 2.45×10⁻⁴), and all had similar directions of effect (**Table 2**). The genomic inflation factor of the PND meta-analysis was 1.07, but the LD Score regression intercept was 0.96, indicating that the inflation was likely attributable to polygenicity rather than confounding.

**Table 2.** Top five linkage disequilibrium-independent loci in the PND GWAS meta-analysis.

|  |  |  |  |  |  |  | PND meta-analysis<br>(this paper) |  | PGC PND<br>(Guintivano et al., 2023) |  | PGC MDD<br>No23andMe, No UKB<br>(Adams et al., 2025) |  |
| --- | --- | --- | --- | --- | --- | --- | --- | --- | --- | --- | --- | --- |
| CHR | SNP | Position | EA | NEA | MAF | Nearest<br>gene | BETA<br>(SE) | P | BETA<br>(SE) | P | BETA<br>(SE) | P |
| 20 | rs6072268 | 39713535 | A | G | 0.157 | *TOP1* | 0.088<br>(0.02) | 6.89x10<sup>-8</sup> | 0.067<br>(0.02) | 1.43x10<sup>-4</sup> | 0.014<br>(0.004) | 2.45x10<sup>-4</sup> |
| 1 | rs7512263 | 96941158 | A | C | 0.337 | *EEF1A1P11/*<br>*RN7SL831P* | 0.068<br>(0.01) | 1.06x10<sup>-7</sup> | 0.07<br>(0.01) | 2.35x10<sup>-7</sup> | 0.007<br>(0.003) | 0.026 |
| 5 | rs13156549 | 139514964 | T | C | 0.228 | *IGIP/*<br>*CTB-131B5.5* | 0.072<br>(0.01) | 7.56x10<sup>-7</sup> | 0.081<br>(0.02) | 3.83x10<sup>-7</sup> | 0.0007<br>(0.003) | 0.827 |
| 6 | rs16882731 | 19811887 | T | C | 0.025 | *RFP1-167F1.2* | 0.188<br>(0.04) | 9.82x10<sup>-7</sup> | 0.192<br>(0.04) | 4.50x10<sup>-6</sup> | 0.007<br>(0.01) | 0.488 |
| 2 | rs12473317 | 52261308 | A | G | 0.231 | *AC007682.1* | -0.070<br>(0.01) | 1.05x10<sup>-7</sup> | -0.071<br>(0.02) | 7.09x10<sup>-6</sup> | -0.002<br>(0.004) | 0.495 |

CHR=chromosome; SNP=Single nucleotide polymorphism; Position=base pair position genome build 37; EA=effect allele; NEA=non-effect allele; MAF=minor allele frequency. PND=perinatal depression, PGC=psychiatric genomics consortium, MDD=major depressive disorder. Results of these SNPs in recent GWAS of PND and MDD are also shown

### SNP heritability and Genetic correlation

The estimated liability scale h^2^_SNP_ of PND in the meta-analysis was 0.089 (SE=0.007), assuming a lifetime risk of 0.10. To assess the shared genetic architecture between PND and other traits, and how they compare with MDD, genetic correlation estimates were computed using LDSC. There was a significant genetic correlation between PND and a recent MDD GWAS(24) of 0.79 (SE=0.03, *P* = 6.97×10^−136^). On average, genetic correlations between PND and MDD with the tested traits were consistent (**Figure 2, Supplementary Table 3**). Both PND and MDD showed negative genetic correlations with age at menopause, progesterone levels, age at menarche, and sleep duration. However, the magnitude of genetic correlation differed between PND and MDD for several traits, with some of these differences being statistically significant (**Supplementary Table 3**). Notably, PND had a higher genetic correlation with anxiety disorders compared to MDD (r_gPND_(SE)= 0.89(0.05) vs r_gMDD_(SE)= 0.77 (0.03), *P*_diff_=0.04). In contrast, MDD exhibited statistically significantly higher positive genetic correlations than PND with cannabis use disorder (*P*_diff_=0.008), bipolar disorder (*P*_diff_=0.03) and ADHD (*P*_diff_=0.03). We also examined the genetic correlation of PND and MDD with a separate female-only MDD GWAS(27). Both traits showed high genetic correlations with female MDD (r_gPND_(SE)= 0.97(0.04), r_gMDD_(SE)= 0.92(0.02)). Although the r_g_ between PND and female MDD was slightly higher than that between MDD and female MDD, this difference was not statistically significant (*P*_diff_=0.24).

**Figure 2.**
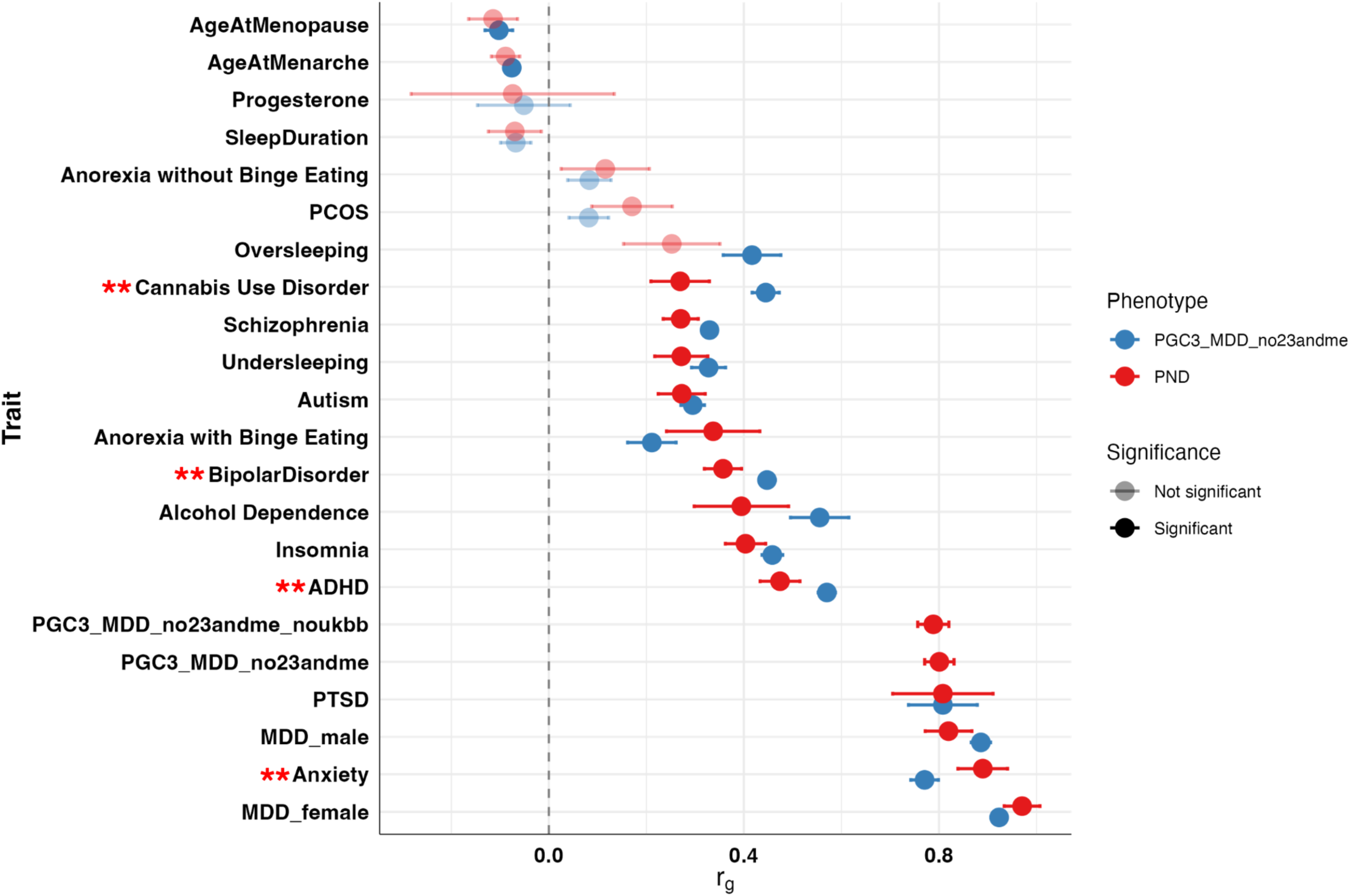
Genetic correlation (rg) between PND, MDD and other traits. Significant rg estimates after multiple testing correction (*P* < 2.27×10^−3^) are shown as bold circles; non-significant estimates are shown as faded circles. Double asterisks (**) indicate traits with significantly different genetic correlations between PND and MDD, Pdiff < 0.05. PGC3_MDD=Wave 3 of the Psychiatric Genomics Consortium summary statistics for MDD, PCOS=Polycystic Ovarian Syndrome, ADHD=Attention-Deficit/Hyperactivity Disorder, PTSD=Post-traumatic Stress Disorder. Details of studies used in **Supplementary Table 2.**

### Gene-based and gene set analyses

The GWAS SNPs were mapped to 19,845 protein-coding genes using MAGMA as implemented in FUMA. None of the MAGMA gene-based associations passed the threshold for Bonferroni correction for multiple testing (*P*=0.05/19845=2.52×10^−6^) (**Supplementary Table 4**). The most significant association was the *ERAL1* gene on chromosome 17 (*P*=6.62×10^−6^).

The MAGMA competitive test of gene sets did not identify any significant associations after Bonferroni correction for multiple testing (*P*=0.05/15,496=3.22×10^−6^). **Supplementary Table 5** provides the top gene sets.

### Tissue expression analysis

Downstream MAGMA expression profiles and competitive pathway analyses were conducted as part of the FUMA pipeline. None of the tissues tested passed the threshold for Bonferroni correction for multiple testing (*P*=0.05/54 tissues=9.26×10^−4^) (**Figure 3**). The highest-ranked tissue-specific expression was observed in brain regions, including the cortex and the nucleus accumbens of the basal ganglia. **Supplementary Table 6** presents the full tissue expression profile results.

**Figure 3.**
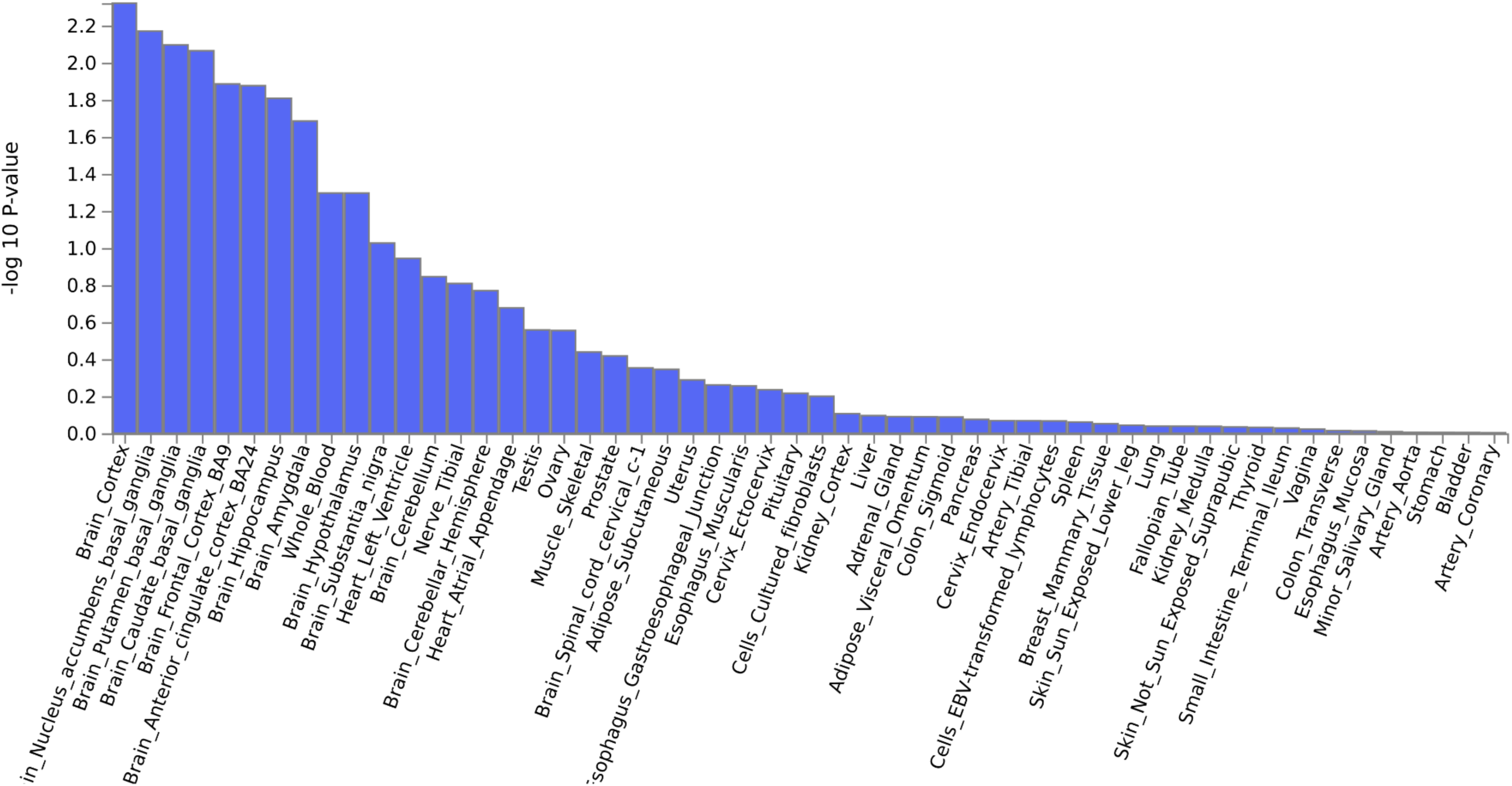
Analysis of tissue enrichment by FUMA using GTEx version 8 (n=54 tissues)

## DISCUSSION

In this study, we present the largest European ancestry meta-analysis of postnatal depression (PND) to date, combining data from UKB and the GLAD Study and a recent PND GWAS from the PGC(10). No genome-wide significant loci were identified. The SNP-based heritability for PND in our study was estimated at 8.9%. While this value falls below the 14% reported in the previous PND GWAS(10), it is within the range of estimates reported for MDD, such as 8.4% in a recent study(24). PND and MDD showed broadly similar genetic correlation patterns with other traits, though some correlations differed significantly in magnitude.

Among the top loci identified in our analysis, the *TOP1* (DNA topoisomerase 1) gene plays a role in DNA transcription and replication and has been previously implicated in neuronal function and stress responses(28), though its role in psychiatric disorders remains underexplored. The tissue expression analysis using GTEx also did not reach statistical significance, but the observation that the highest ranked enrichment in brain regions, such as the cortex and the nucleus accumbens of the basal ganglia, suggests these areas may play a role in the biological mechanisms underlying PND. Larger sample sizes in genetic studies of PND may enable clarification on genome-wide loci and tissue expression.

The lower heritability estimate compared to the previous PND GWAS may partly reflect differences in phenotype definition. While most cohorts in the earlier study used the EPDS to define PND, providing a stricter and more clinically grounded definition of the phenotype, the UKB and GLAD samples relied primarily on self-report. Despite this difference in case definition, our findings align with the earlier study in showing that PND is more heritable than MDD overall (h^2^_SNP_ 0.089 PND vs 0.077 MDD(24)), supporting the notion that PND may have a stronger genetic component(3).

We used genetic correlation to assess whether depression during pregnancy and the postnatal period (PND) is genetically distinct from overall MDD. First, we observed high genetic correlation between PND and female MDD (r_g_=0.97), which was only marginally higher than the correlation between MDD and female MDD (r_g_=0.92). While this is expected, given that our analysis focused on female PND cases, it raises questions about whether the genetic overlap with female MDD is specific to the perinatal period or reflects broader shared liability. Future studies will be needed to disentangle this. Second, PND showed a significantly higher genetic correlation with anxiety disorders than MDD. One plausible explanation is that the EPDS, used in the majority of PND studies in the meta-analysis, captures symptoms of anxiety alongside depression. Indeed, previous studies have questioned the specificity of the EPDS for depression, suggesting that it may reflect a broader affective profile(29,30). Third, the significantly lower genetic correlations of PND with bipolar disorder, cannabis use disorder and ADHD compared to MDD suggest that the underlying genetic variants contribute more to MDD and anxiety than other psychiatric disorders. However, this result could be a function of the lower sample sizes of PND (n=97,583) compared to MDD (n=752,566), as the increase in sample size in larger studies may increase phenotypic heterogeneity(31).

This study has several limitations that should be considered when interpreting the findings, while also laying the groundwork for future research. First, the sample size was not large enough to detect genome-wide significant loci, though efforts to increase sample sizes are ongoing. Second, additional analyses in ancestrally diverse datasets are needed to ensure findings are broadly applicable. Greater diversity in genetic studies has been shown to improve power to detect risk loci that may be missed in European-only samples(24). Third, the timing of the PND phenotype requires careful attention. While often used interchangeably with postpartum depression (PPD), PND can include both the pregnancy period and up to 12 months after birth(32). Our meta-analysis included studies with varying and sometimes overlapping definitions. Clarifying the timing of onset is essential to better understand the aetiology of PND and how it differs from MDD. Lastly, selection biases in GLAD+ and UKB warrant consideration. UKB participants are older (median age 58 years in this study) and more socioeconomically advantaged than the general population(9), which may influence both the likelihood of participating in mental health assessments and the accuracy of retrospective reporting of perinatal depression. Additionally, GLAD+ cases tend to represent individuals with more severe or recurrent depression, whereas UKB cases are generally milder(33). These differences in age, socioeconomic background, and clinical severity may influence phenotype distributions and limit the comparability of effect estimates across cohorts.

In summary, this study adds new data from UKB and GLAD to the existing GWAS of PND. Genetic correlation results suggest there may be distinct genetic factors differentiating PND from MDD. The absence of genome-wide significant findings and the limited polygenic score transferability highlight the need for larger and more ancestrally diverse studies.

## Data Availability

All data produced in the present study are available upon reasonable request to the authors

## ACKNOWLEDGEMENTS

We thank the GLAD+ Studies, NIHR BioResource, and UK Biobank volunteers for their participation and gratefully acknowledge the contributions of NIHR BioResource centres, NHS Trusts, UK Biobank, and staff. We also thank the National Institute for Health & Care Research (NIHR), NHS Blood and Transplant, and Health Data Research UK as part of the Digital Innovation Hub Programme.

This study represents independent research funded by the NIHR Biomedical Research Centre at South London and Maudsley NHS Foundation Trust and King’s College London. Further information is available at https://www.maudsleybrc.nihr.ac.uk/facilities/bioresource/. The views expressed are those of the authors and do not necessarily reflect those of the NHS, NIHR, HSC R&D Division, King’s College London, or the Department of Health and Social Care.

## CONFLICT OF INTEREST

Prof Breen has received honoraria, research or conference grants and consulting fees from Illumina, Otsuka, and COMPASS Pathfinder Ltd. Dr Davis has received a research grant from SPIN-Dementia Network+.

## FUNDING

This work was supported by the National Institute for Health and Care Research (NIHR) BioResource [RG94028, RG85445], NIHR Biomedical Research Centre [ISBRC-1215-20018], HSC R&D Division, Public Health Agency [COM/5516/18], MRC Mental Health Data Pathfinder Award (MC_PC_17,217), and the National Centre for Mental Health funding through Health and Care Research Wales. JGM acknowledges funding from Wellcome DepGenAfrica grant number 223165/Z/21/Z.

## SUPPLEMENTARY INFORMATION

Supplementary information is available at MP’s website

## Notes

### Author Declarations

Ethics committee/Institutional Review Board of King's College London gave ethical approval for this work

